# Exploring Beauty Product Accessibility for Individuals with Upper Extremity Disabilities

**DOI:** 10.1101/2024.01.29.24301948

**Authors:** Niko Fullmer, Hannah Cone, Jeanette Gumarang, Emily Kieffer, Soyoung Esther Bae, Emily R Rosario

**Affiliations:** Casa Colina Research Institute; Modified Independent

**Keywords:** Accessibility, Upper Extremity Disabilities, Neurologic conditions, Activities of daily Living, Satisfaction

## Abstract

**Objective:** This study explores the accessibility of beauty products for individuals with upper extremity disabilities.

**Methods:** Participants with varied upper extremity impairments used Rare Beauty makeup products over two weeks. Assessments of hand function and questionnaires evaluated usability and satisfaction.

**Results:** Product features like bottle shape, cap design, and texture significantly influenced usability for those with hand and arm impairments. Notably, individuals with fine motor skill impairments reported easier gripping with larger bottle sizes and ease of opening with cap enhancements. The unique product shape and enhanced caps were also found to be more accessible for participants across all ability levels. Overall, the products were well-received, with most participants finding them comfortable and user-friendly.

**Conclusions:** The study emphasizes the need for inclusive design in the beauty industry, catering to those with upper extremity disabilities. It reveals the importance of ergonomic and adaptable product features to enhance usability and accessibility.

**Plain Language summary:** This study investigated the use of beauty products by people with upper extremity disabilities. Participants tried Rare Beauty makeup for two weeks and provided feedback. Results showed that product design, like bottle shape and cap ease, significantly impacts usability for those with hand and arm function impairments. Key findings included better grip on certain products for those with fine motor skill challenges. Overall, participants found the products comfortable and easy to use, highlighting the importance of inclusive design in beauty products for people with disabilities.

## Introduction

Hand function and upper extremity capabilities play a crucial role in our daily lives, profoundly impacting activities of daily living (ADLs), encompassing a broad spectrum from basic self-care like eating and dressing to more complex activities such as using technology or engaging in hobbies (D. Kim, 2016; Yamamoto et al., 2022; Yoo & Park, 2015). The upper extremities, including hands, arms, and shoulders, facilitate a diverse range of movements, essential for interacting with our environment and maintaining independence (D. Kim, 2016; Pollock et al., 2014; Synek et al., 2023). This relationship between upper extremity function and ADLs is a vital area of study in fields such as occupational therapy and rehabilitation medicine (Alsubiheen et al., 2022; Pollock et al., 2014). Impairments in these areas may be due to injury, neurological disorders, congenital disorders, or degenerative conditions. For example, conditions like stroke, spinal cord injury, or arthritis can limit the strength, range of motion, and coordination of the upper extremities, challenging everyday tasks such as dressing, eating, or even using digital devices (D. Kim, 2016; Mano et al., 2018; Richards & Pohl, 1999; Yamamoto et al., 2022). This leads to a critical need for therapeutic interventions and adaptive strategies to enhance or maintain functional independence.

The realm of beauty products, a vital part of daily life for many, often overlooks the unique needs of individuals with upper extremity disabilities (Nast, 2021; Zhang et al., 2020). This oversight impacts not only the usability of these products but also the autonomy and self-esteem of this population (Entwistle et al., 2010; Wu & Chao, 2023). While there has been some advancement in accessible design across various sectors, beauty products specifically designed for individuals with upper extremity disabilities or with adaptations for this population are still scarce (Gregg, 2023; S. Kim, 2022). Beauty items, ranging from cosmetics to skincare, are predominantly designed with standard packaging and applicators that lack features that address the specific challenges faced by those with limited mobility or dexterity, creating a significant design gap. The lack of consideration for diverse physical abilities can result in barriers to accessing and using these products effectively (Gallagher, 2021; Nast, 2021).

Oversight in product design often leads to difficulties in gripping, opening, or maneuvering beauty items, thereby limiting the independence of individuals with upper extremity disabilities (Gregg, 2023; Gallagher, 2021; Nast, 2021). Products that are not user-friendly can contribute to frustration and may even discourage participation in beauty routines. The inability to access and use beauty products can significantly impact self-esteem, as personal grooming and appearance are essential aspects of self-expression. Accessible beauty products that consider factors such as ease of use, application and ergonomic design, on the other hand, empower individuals with disabilities to maintain a sense of autonomy in their daily lives (Bido, 2023; Entwistle et al., 2010; Toscano, 2023a, 2023b). Enabling independence in the use of beauty products not only contributes to a positive self-image but also enhances overall well-being.

The beauty industry is witnessing a rising awareness of disability inclusivity, with a handful of recognized brands, like Rare Beauty and Guide Beauty, taking the lead. Notably these brands’ founders are open about their lived experiences with disability. But the journey towards broader inclusivity that deliberately meets the needs of consumers with disabilities is still in its nascent stages despite clear user feedback on the challenges faced by individuals with disabilities (Camero, 2023; Gregg, 2023). There is a lack of commitment in terms of attention and resources to create products inclusive of individuals with disabilities. Furthermore, there is a significant research gap in addressing the accessibility of beauty products for these individuals. Understanding their unique needs in greater depth is crucial for crafting effective, inclusive solutions. Overcoming this gap necessitates a more proactive stance in integrating a variety of user experiences into design processes. Research in the field of accessible design underscores the importance of developing inclusive products in the cosmetics and personal care industry, advocating for the incorporation of ergonomic designs and adaptive features to better serve individuals with upper extremity impairments. This study, therefore, aimed to bridge this gap by investigating and enhancing the accessibility of beauty products for individuals with upper extremity disabilities. The significance of this research lies not only in its potential to influence product design but also in its implications for occupational therapy, where the focus on enhancing daily living skills and independence is paramount. Enhancing the accessibility of beauty products directly contributes to improving the overall quality of life for individuals with upper extremity disabilities. This research recognizes the role that personal grooming and appearance impact well-being and seeks to positively influence these aspects of daily life. Occupational therapists can utilize insights from this research to tailor interventions that address specific challenges related to beauty routines, ultimately enhancing their clients’ overall independence.

## Methods

### Study Design

A single-group pre-test/post-test design was used to determine the accessibility of seven beauty products from the Rare Beauty line. Participants, comprising of individuals aged 18 and above with various upper extremity disabilities, completed pre-test questionnaires and a functional assessment to establish a baseline of their demographics, disability characteristics, and prior experiences with beauty products. This was followed by a trial period where participants used a selection of beauty products specifically chosen for their potential accessibility features. Participants completed post-test questionnaires to assess any changes in their perceptions and experiences with the products.

### Participants

The participants included individuals with a range of upper extremity impairments. Efforts were made to ensure diversity in terms of the type and severity of disabilities, as well as previous experiences with beauty products. The inclusion criteria focused on adults with documented upper extremity disabilities, while exclusion criteria included individuals under 18 and those with cognitive impairments affecting their ability to consent or participate effectively.

### Procedure

#### Pre-test Assessment

As part of the initial evaluation, participants underwent grip strength testing using a hand-held dynamometer and the Action Research Arm Test. These assessments provided insights into their gross motor skills, vital for tasks like lifting, holding, and manipulating objects, and their fine motor skills, which are crucial for precise grasping and dexterous hand movements. Participants were categorized into different impairment levels based on established norms and cutoffs for these standardized outcome measurements. Participants also completed pre-test questionnaires to collect detailed information about their demographics, disability characteristics, and previous experiences with beauty products.

#### Trial Period

During the trial period, participants were given various Rare Beauty makeup products, selected for their potential accessibility. They were instructed to incorporate these products into their normal beauty routines over a two-week span. The trial involved testing seven Rare Beauty products, with three products used in the first week and four in the second. After each week, participants completed questionnaires to provide feedback on their experience with these products.

#### Post-trial Assessment

After the trial period, participants completed post-test questionnaires to reflect on their experiences, emphasizing satisfaction, usability, and accessibility. Additionally, they assessed the beauty products on ease of grip, maneuverability, packaging, and applicators, noting challenges and possible improvements. The study’s outcome measures included qualitative evaluations of usability, satisfaction, and barriers to using standard beauty products, along with quantitative ratings for ease of use, comfort, and overall product satisfaction. Time taken for product use and any required assistance were also recorded.

### Analysis

The analysis included both quantitative and qualitative methodologies. Qualitative data from participant feedback were analyzed thematically to identify patterns and insights related to the accessibility and usability of the beauty products. ChatGPT was used to assist with this analysis. Quantitative data included post-trial questionnaires as well as grip, tip, and key strength scores, and ARAT results. Chi-square analyses were performed using JMP software to examine potential relationships between product design elements and the participants’ impairment level.

### Ethical Considerations

The study adhered to ethical standards, with approval from the Institutional Review Board. Participants provided informed consent and were assured of confidentiality and the right to withdraw at any time. Data were securely stored and accessible only to the research team.

## Results

The study consisted of 57 individuals with a range of upper extremity disabilities, including stroke, TBI, SCI, and other neurologic and orthopedic conditions. The average age was 43.8 years, with a diverse distribution in terms of ethnicity, education, and disability levels in both gross and fine motor skills (Table 1). Participants with upper extremity disabilities reported various challenges during their makeup routine on the pre-test questionnaire. Physical difficulties included cramps, loss of sensation, and coordination issues, making it hard to maintain arm positions. Fine motor skill challenges were most pronounced with small makeup items like eyeliner and eyeshadow, requiring precise application. Bimanual tasks such as handling and opening containers was another challenging area, requiring the use of compensatory strategies, such as utilizing the mouth for assistance. Visual and cognitive impairments also complicated the makeup process, affecting routine memory and product application. When asked about specific product difficulties, eyeshadow, eyeliner, and mascara were frequently cited as challenging due to their size and the precision needed for application. Narrow and small products, such as concealers and eyeshadows in stick or pen form, were identified as problematic, as were items with twist-off caps. Participants also found compact containers and pressed powder products difficult to handle. While some found pump dispensers more accessible, others struggled with them, while spray products posed challenges for some participants as well.

**Table 1.**
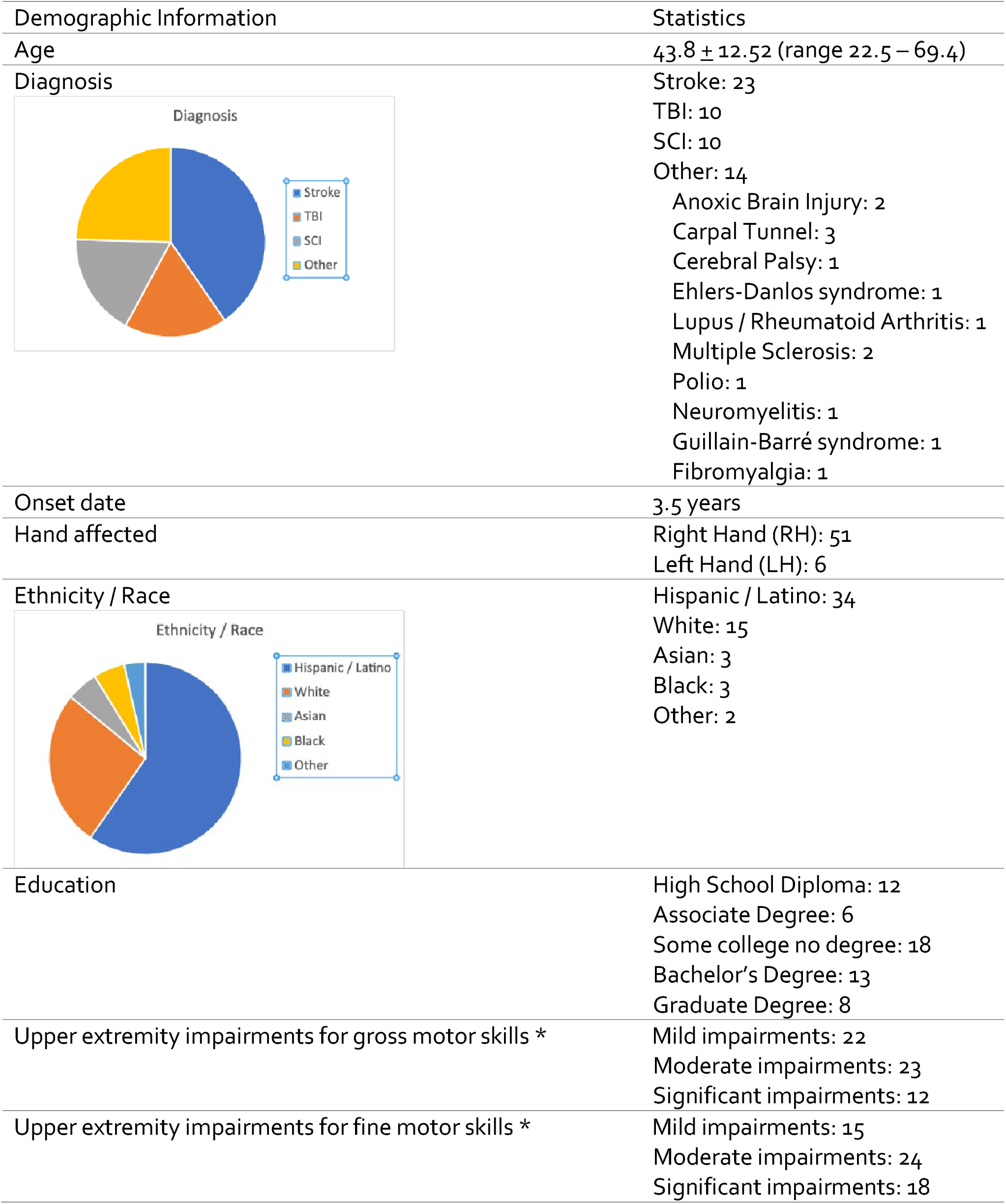
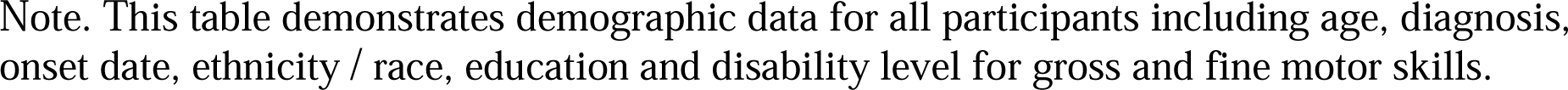
Participant Demographics.

| Demographic Information | Statistics |
| --- | --- |
| Age | 43.8 ± 12.52 (range 22.5 – 69.4) |
| Diagnosis | Stroke: 23<br>TBI: 10<br>SCI: 10<br>Other: 14<br>Anoxic Brain Injury: 2<br>Carpal Tunnel: 3<br>Cerebral Palsy: 1<br>Ehlers-Danlos syndrome: 1<br>Lupus / Rheumatoid Arthritis: 1<br>Multiple Sclerosis: 2<br>Polio: 1<br>Neuromyelitis: 1<br>Guillain-Barré syndrome: 1<br>Fibromyalgia: 1 |
| Onset date | 3.5 years |
| Hand affected | Right Hand (RH): 51<br>Left Hand (LH): 6 |
| Ethnicity / Race | Hispanic / Latino: 34<br>White: 15<br>Asian: 3<br>Black: 3<br>Other: 2 |
| Education | High School Diploma: 12<br>Associate Degree: 6<br>Some college no degree: 18<br>Bachelor's Degree: 13<br>Graduate Degree: 8 |
| Upper extremity impairments for gross motor skills * | Mild impairments: 22<br>Moderate impairments: 23<br>Significant impairments: 12 |
| Upper extremity impairments for fine motor skills * | Mild impairments: 15<br>Moderate impairments: 24<br>Significant impairments: 18 |

Note. This table demonstrates demographic data for all participants including age, diagnosis, onset date, ethnicity / race, education and disability level for gross and fine motor skills.

Participants were also asked about accessible beauty products in the pre-test questionnaires. Participants indicated that items with pump dispensers, larger containers and brushes, as well as cream-based products were easier to use. Unique designs such as slide packaging and products with thick-handled wands or applicators were also called out for improved accessibility. Preferred product packaging included easy-to-grip, matte, glass materials and secure closure mechanisms. Difficulties were noted with opening certain types of packaging, such as lipstick and moisturizers. Suggestions for improvement included longer applicator wands, changes in packaging materials for easier grip, and design modifications to accommodate specific medical conditions.

Participant feedback from the trial period underscores that the design features of beauty products, such as packaging ergonomics—encompassing bottle shape, cap design, and texture— noticeably influence their accessibility and usability. Notably, a vast majority of participants (77-96%) praised the design of the products’ containers for their comfort on hands and wrists and their ability to minimize dropping or slipping compared to traditional pump/squeeze bottles. Over 80% of participants found the cap of several products easier to hold and in many cases easier to open. Additionally, over 85% of users agreed that the product applicator shape facilitated easy and effective application, while the blendability and consistency of the product formulas were positively rated by 80% or more participants, improving and enhancing the experience of even product application (data for all products in Tables 2 – 3 and Supplemental Tables 6-10).

**Table 2.** User Feedback on Liquid Touch Weightless Foundation.

| Question | Statistic | Count | Percent |
| --- | --- | --- | --- |
| Does the finish of the bottle allow you to grip it more firmly and prevent it from slipping? | Agree | 47 | 84% |
|  | Neutral | 6 | 11% |
|  | Disagree | 3 | 5% |
| Does the applicator wand easily pull out from the bottle? | Agree | 48 | 86% |
|  | Neutral | 3 | 5% |
|  | Disagree | 5 | 9% |
| Does the cap make the applicator easier to grip? | Agree | 49 | 88% |
|  | Neutral | 6 | 11% |
|  | Disagree | 1 | 1% |
| Does the cap make the product easier to open? | Agree | 37 | 66% |
|  | Neutral | 12 | 21% |
|  | Disagree | 7 | 13% |
| Does the cap make it easier to apply the product to your face? | Agree | 43 | 77% |
|  | Neutral | 11 | 20% |
|  | Disagree | 2 | 3% |
| Does the shape of the bottle allow you to comfortably hold the bottle while you open the foundation? | Agree | 51 | 92% |
|  | Neutral | 2 | 3% |
|  | Disagree | 3 | 5% |
| Does the shape of the applicator make it easier to apply the product than a foundation pump/squeeze bottle? | Agree | 51 | 92% |
|  | Neutral | 2 | 3% |
|  | Disagree | 3 | 5% |
| Does the fluidity of the formula allow you to blend the product easier? | Agree | 45 | 80% |
|  | Neutral | 7 | 13% |
|  | Disagree | 4 | 7% |
| Does the consistency of the formula allow for enough time for an even application? | Agree | 47 | 84% |
|  | Neutral | 6 | 11% |
|  | Disagree | 3 | 5% |
| Is it comfortable for your hands and wrists to use this product? | Agree | 54 | 96% |
|  | Disagree | 2 | 4% |

Note. This table demonstrates the number and percent of participants who agreed, were neutral, or disagreed with each of the above questions following a 1-week trial period with the product.

**Table 3.** User Feedback on Positive Light Liquid Luminizer.

| Question | Statistic | Count | Percent |
| --- | --- | --- | --- |
| Does the size of the bottle allow you to grip it more securely? | Agree | 45 | 80% |
|  | Neutral | 7 | 13% |
|  | Disagree | 4 | 7% |
| Does the applicator wand easily pull out from the bottle? | Agree | 49 | 88% |
|  | Neutral | 2 | 3% |
|  | Disagree | 5 | 9% |
| Does the cap make the product easier to grip? | Agree | 48 | 86% |
|  | Neutral | 5 | 9% |
|  | Disagree | 3 | 5% |
| Does the cap make the product easier to open? | Agree | 41 | 73% |
|  | Neutral | 8 | 14% |
|  | Disagree | 7 | 13% |
| Does the cap make it easier to apply the product to your face? | Agree | 46 | 82% |
|  | Neutral | 8 | 14% |
|  | Disagree | 2 | 4% |
| Does the shape of the bottle allow you to comfortably hold the bottle while you open the liquid luminizer? | Agree | 50 | 90% |
|  | Neutral | 3 | 5% |
|  | Disagree | 3 | 5% |
| Does the shape of the applicator make it easier to apply the product? | Agree | 49 | 88% |
|  | Neutral | 5 | 9% |
|  | Disagree | 2 | 3% |
| Does the fluidity of the formula allow you to blend the product easier? | Agree | 49 | 88% |
|  | Neutral | 5 | 9% |
|  | Disagree | 2 | 3% |
| Does the consistency of the formula allow for enough time for an even application? | Agree | 48 | 86% |
|  | Neutral | 7 | 13% |
|  | Disagree | 1 | 1% |
| Is it comfortable for your hands and wrists to use this product? | Agree | 50 | 89% |
|  | Neutral | 5 | 9% |
|  | Disagree | 1 | 2% |

Note. This table demonstrates the number and percent of participants who agreed, were neutral, or disagreed with each of the above questions following a 1-week trial period with the product.

A more detailed examination of these product design elements using chi-square analysis found significant differences in user experience based on level of upper extremity impairments. Participants with mild to moderate fine motor impairments, for example, had a significant preference for the grip of the Liquid Touch Weightless Foundation bottle (Chi-sq = 9.3*), pointing to how tailored product design can markedly improve usability for individuals with different dexterity levels. Similarly, the Positive Light Liquid Luminizer’s bottle size and the ease with which the wand could be removed were significant aspects of product satisfaction for individuals with more pronounced fine motor impairments (Chi-sq = 10.54* and 9.56*, respectively). For the Soft Pinch Tinted Lip Oil, the texture of the bottle and the design of the handle significantly impacted the comfort and ease of application for participants with both gross motor and fine motor impairments (Chi-sq = 12.75*). Additionally, the cap of the Kind Words Matte Lipstick was noted for its comfortable grip, especially by users with fine motor impairments who found it significantly easier to manage (Chi-sq = 9.59*). The overall feedback from the study suggests that while all the products tested were comfortable and easy to use across different levels of disability, the highest comfort levels were reported by individuals with more severe fine motor impairments (data for all products in Tables 4 – 5 and Supplemental Tables 11-15).

**Table 4.**
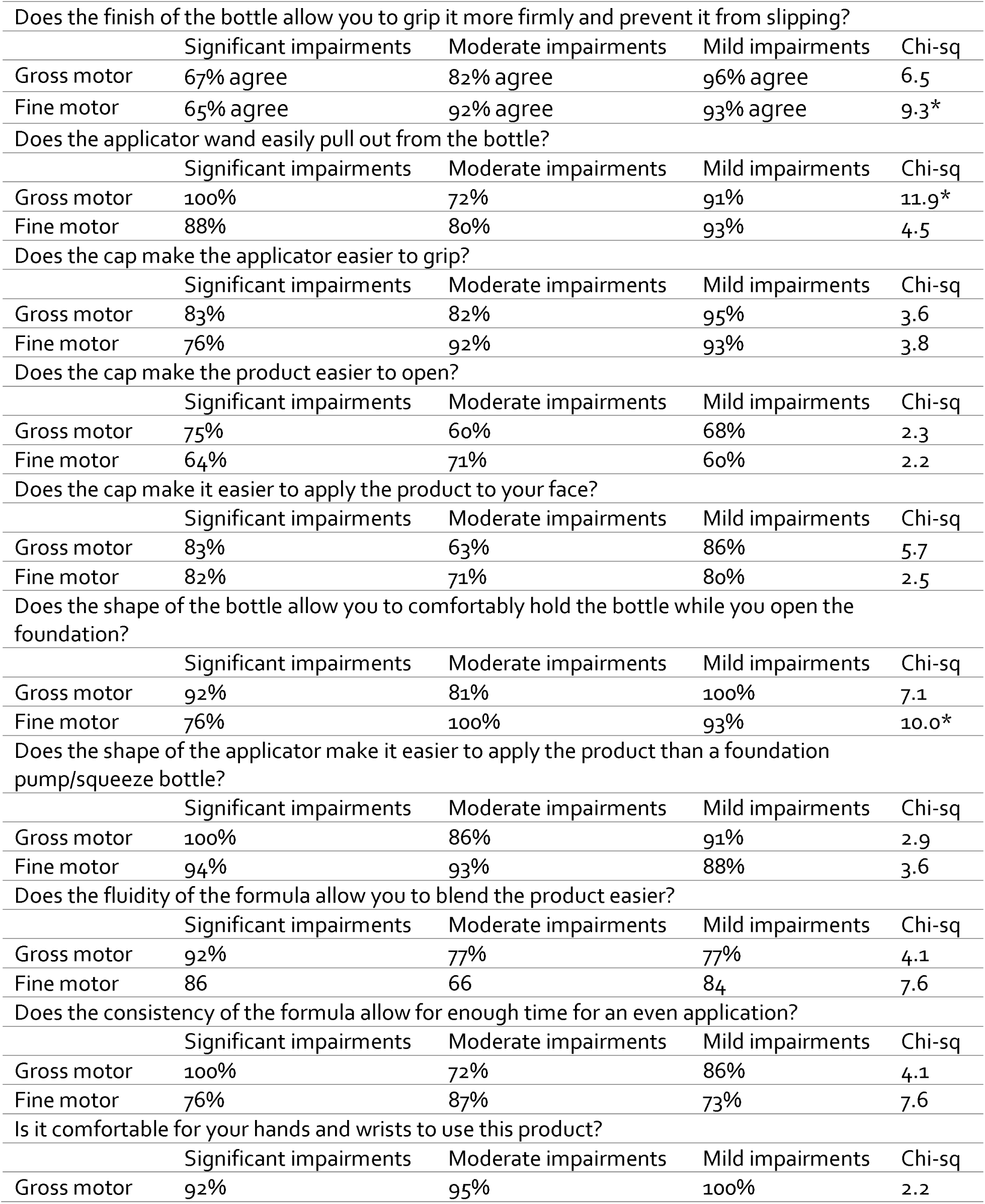
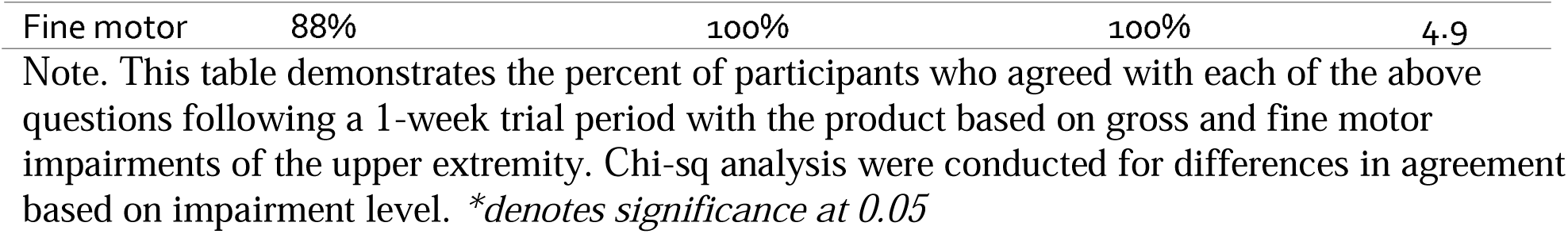
Percent of positive feedback on Liquid Touch Weightless Foundation by Ability Level.

**Table 5.**
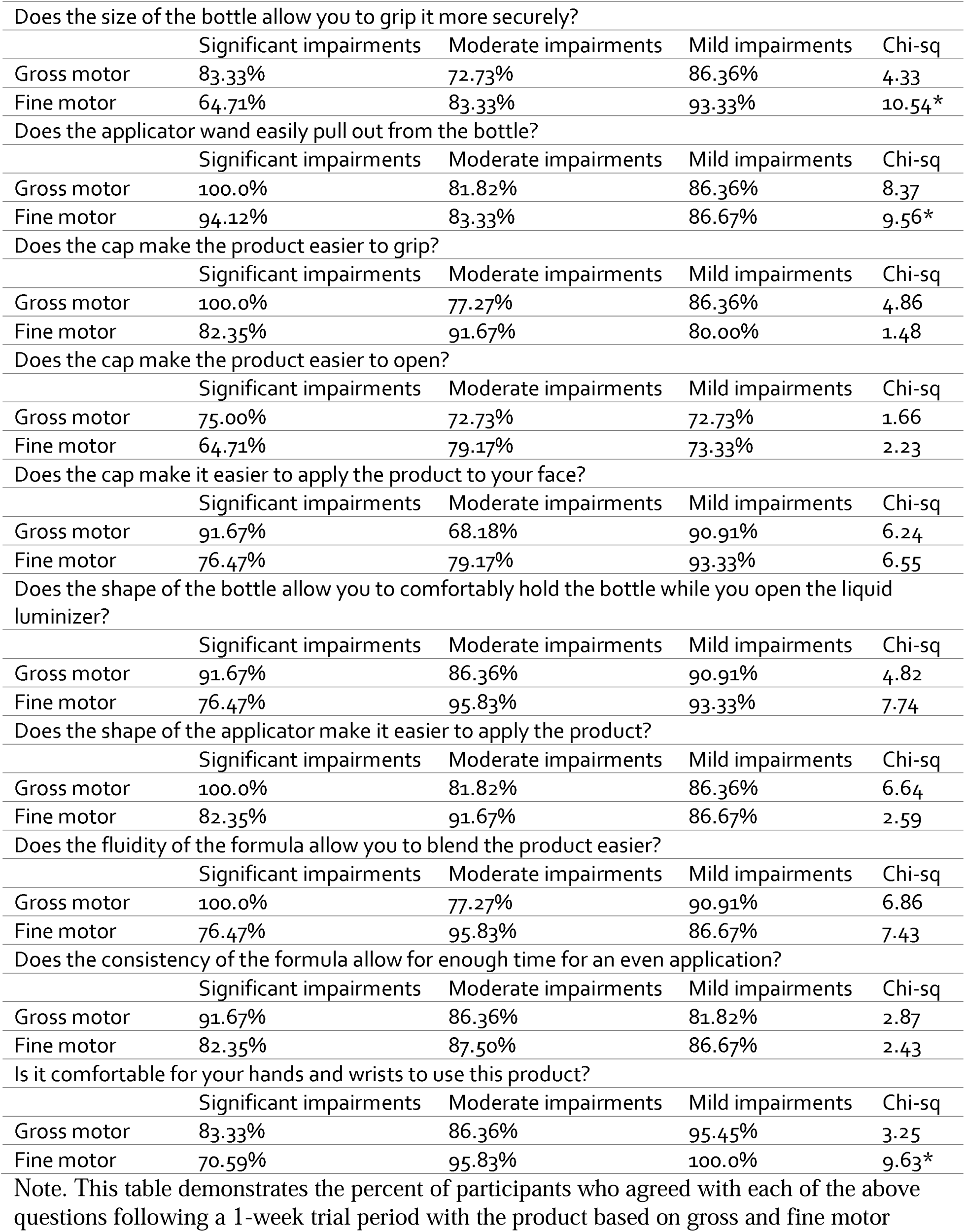
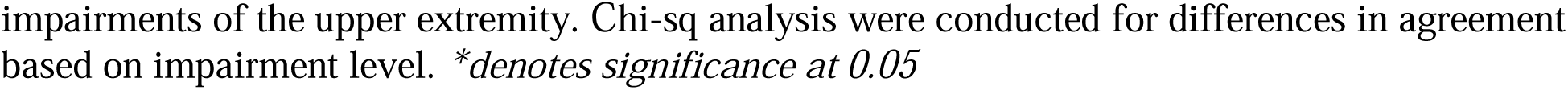
Percent of positive feedback on Positive Light Liquid Luminizer by Ability level.

|  |  |  |  |  |
| --- | --- | --- | --- | --- |
| Does the size of the bottle allow you to grip it more securely? |  |  |  |  |
|  | Significant impairments | Moderate impairments | Mild impairments | Chi-sq |
| Gross motor | 83.33% | 72.73% | 86.36% | 4.33 |
| Fine motor | 64.71% | 83.33% | 93.33% | 10.54* |
| Does the applicator wand easily pull out from the bottle? |  |  |  |  |
|  | Significant impairments | Moderate impairments | Mild impairments | Chi-sq |
| Gross motor | 100.0% | 81.82% | 86.36% | 8.37 |
| Fine motor | 94.12% | 83.33% | 86.67% | 9.56* |
| Does the cap make the product easier to grip? |  |  |  |  |
|  | Significant impairments | Moderate impairments | Mild impairments | Chi-sq |
| Gross motor | 100.0% | 77.27% | 86.36% | 4.86 |
| Fine motor | 82.35% | 91.67% | 80.00% | 1.48 |
| Does the cap make the product easier to open? |  |  |  |  |
|  | Significant impairments | Moderate impairments | Mild impairments | Chi-sq |
| Gross motor | 75.00% | 72.73% | 72.73% | 1.66 |
| Fine motor | 64.71% | 79.17% | 73.33% | 2.23 |
| Does the cap make it easier to apply the product to your face? |  |  |  |  |
|  | Significant impairments | Moderate impairments | Mild impairments | Chi-sq |
| Gross motor | 91.67% | 68.18% | 90.91% | 6.24 |
| Fine motor | 76.47% | 79.17% | 93.33% | 6.55 |
| Does the shape of the bottle allow you to comfortably hold the bottle while you open the liquid luminizer? |  |  |  |  |
|  | Significant impairments | Moderate impairments | Mild impairments | Chi-sq |
| Gross motor | 91.67% | 86.36% | 90.91% | 4.82 |
| Fine motor | 76.47% | 95.83% | 93.33% | 7.74 |
| Does the shape of the applicator make it easier to apply the product? |  |  |  |  |
|  | Significant impairments | Moderate impairments | Mild impairments | Chi-sq |
| Gross motor | 100.0% | 81.82% | 86.36% | 6.64 |
| Fine motor | 82.35% | 91.67% | 86.67% | 2.59 |
| Does the fluidity of the formula allow you to blend the product easier? |  |  |  |  |
|  | Significant impairments | Moderate impairments | Mild impairments | Chi-sq |
| Gross motor | 100.0% | 77.27% | 90.91% | 6.86 |
| Fine motor | 76.47% | 95.83% | 86.67% | 7.43 |
| Does the consistency of the formula allow for enough time for an even application? |  |  |  |  |
|  | Significant impairments | Moderate impairments | Mild impairments | Chi-sq |
| Gross motor | 91.67% | 86.36% | 81.82% | 2.87 |
| Fine motor | 82.35% | 87.50% | 86.67% | 2.43 |
| Is it comfortable for your hands and wrists to use this product? |  |  |  |  |
|  | Significant impairments | Moderate impairments | Mild impairments | Chi-sq |
| Gross motor | 83.33% | 86.36% | 95.45% | 3.25 |
| Fine motor | 70.59% | 95.83% | 100.0% | 9.63* |

Note. This table demonstrates the percent of participants who agreed with each of the above questions following a 1-week trial period with the product based on gross and fine motor
impairments of the upper extremity. Chi-sq analysis were conducted for differences in agreement based on impairment level. *\*denotes significance at 0.05*

## Discussion

The goal of this study was to evaluate the potential accessibility of various Rare Beauty makeup products for individuals with upper extremity disabilities. The findings emphasize the importance of inclusivity in product design, making these beauty products more accessible and user-friendly for a wider range of individuals. The study’s outcome measures included qualitative evaluations of usability, satisfaction, and barriers to using standard beauty products, along with quantitative ratings for ease of use, comfort, and overall product satisfaction. The data highlighted the significance of product design in catering to the needs of individuals with upper extremity disabilities, considering their motor skill abilities. Ease of handling, cap and cap removal, product application, fluidity and consistency, and overall comfort play essential roles in determining the user experience for these products. The study’s findings revealed that the design of beauty products significantly influences their accessibility and usability for individuals with varying motor skill abilities. For instance, the design of the Liquid Touch Weightless Foundation was well-received, with users appreciating the finish of the bottle, ease of applicator wand usage, and the cap’s grip, making it comfortable for their hands and wrists. Similarly, the Positive Light Liquid Luminizer, Soft Pinch Tinted Lip Oil, Liquid Touch Brightening Concealer, Kind Words Matte Lipstick, Soft Pinch Liquid Blush, and Positive Light Tinted Moisturizer were all positively rated for their design elements, contributing to a comfortable and efficient user experience.

Globally, it is estimated that over 1 billion people, or 15% of the world’s population, live wit some form of disability and that 80% of disabilities are acquired between the ages of 18 and 64, as reported by the World Health Organization (*Factsheet on Persons with Disabilities | United Nations Enable*; *WHO*) This demographic represents a significant portion of the Global population, including a substantial part of the adult population with disposable income (*Diverse Perspectives*).

The disability community, which is the largest minority group in many countries including the United States, stands out as it is the only group that anyone can join at any time due to the unpredictable onset of disabilities (*Factsheet on Persons with Disabilities United Nations Enable*; *WHO*). This study underscored the critical role of motor abilities in the accessibility and usability of beauty products, highlighting the necessity for designs that accommodate a broad spectrum of functional capabilities. The results indicate that while all tested products were found to be comfortable and user-friendly for people with varying levels of disability, those with more severe fine motor impairments experienced the greatest comfort. This finding emphasizes the importance of continuous innovation in product design to address the diverse requirements of all consumers, with a particular focus on individuals with upper extremity disabilities.

### Study limitations

While the study results were encouraging there were several potential limitations that must be considered. The study included individuals with a range of upper extremity disabilities, however, the sample size may not fully capture the diversity of experiences and needs within this population. A larger and more diverse sample could provide a more comprehensive understanding of the accessibility and usability of beauty products for individuals with upper extremity disabilities. Additionally, the study focused specifically on Rare Beauty makeup products and did not include a control group using standard beauty products, which may limit the generalizability and comparability of the findings to other beauty product brands. Different brands may have varying product designs and packaging, which could impact the accessibility and usability for individuals with upper extremity disabilities. Therefore, the findings should be interpreted within the context of the specific products tested. Despite these limitations, the study’s findings offer valuable insights into the importance of product design elements in improving the user experience for individuals with upper extremity disabilities. The positive feedback on the design of the Rare Beauty products highlights the potential impact of inclusive product design on enhancing accessibility and usability for this population. However, future research, as well as a broader range of beauty product brands, could further advance our understanding of accessible beauty products for individuals with upper extremity disabilities.

### Conclusions

In conclusion, the study’s findings provide valuable insights into the accessibility of beauty products for individuals with upper extremity disabilities, emphasizing the importance of inclusive product design. The research highlights the significance of product packaging, including bottle shape, cap design, texture, and size, in influencing the usability of these beauty products for individuals with varying motor skill abilities. The study’s outcomes contribute to the ongoing efforts to make beauty products more accessible and user-friendly for individuals with upper extremity disabilities, ultimately enhancing their overall quality of life and promoting inclusivity in the beauty industry.

## Data Availability

All data produced in the present study are available upon reasonable request to the authors

## Acknowledgements

We would like to thank Rare Beauty for their generous funding and support of this research project. Their commitment to inclusivity and accessibility in beauty products has been instrumental in advancing our study. Additionally, we wish to express our sincere thanks to the CEO and Board of Directors at Casa Colina Hospital and Centers for Health Care for their support of the Casa Colina Research Institute. We would like to express our gratitude to the clinicians and volunteers who assisted with this study. Finally, we acknowledge the use of ChatGPT for its assistance in summarizing and analyzing our dataset. This technology facilitated our research process and helped us derive meaningful insights from our data.

## Declaration of Conflicting Interests

We wish to disclose that our research, titled “***Exploring Beauty Product Accessibility for Individuals with Upper Extremity Disabilities***,” received funding from Rare Beauty. The brand provided financial support for the materials and resources necessary to conduct the study, including the beauty products tested by participants.

Despite this financial contribution, Rare Beauty held no influence over the study’s design, data collection, analysis, or interpretation. Our findings and conclusions were drawn independently, and Rare Beauty had no role in the writing of the manuscript or the decision to submit the paper for publication.

Each author has contributed significantly to the work and maintained academic and research integrity throughout the study. We have conducted our research under the ethical guidelines provided by our institutional review board, which emphasizes the importance of transparency and objectivity in our scientific inquiry.

We declare this conflict of interest to maintain the transparency and uphold the integrity of the research process and the dissemination of our findings.

## Research ethics section and patient consent

The research presented in this manuscript was conducted with the approval of the Casa Colina Hospital Institutional Review Board (IRB) (FWA0006711), adhering to the highest ethical standards in research involving human subjects. The procedures followed were in accordance with the ethical standards of the Casa Colina Hospital IRB and consistent with the revised (2000) Helsinki Declaration, as detailed on the World Medical Association website.

## References

Alsubiheen, A. M., Choi, W., Yu, W., & Lee, H. (2022). The Effect of Task-Oriented Activities Training on Upper-Limb Function, Daily Activities, and Quality of Life in Chronic Stroke Patients: A Randomized Controlled Trial. International Journal of Environmental Research and Public Health, 19(21), 14125. 10.3390/ijerph192114125

Bido, T. (2023, May 25). 7 Products That Make Beauty More Accessible. NewBeauty. https://www.newbeauty.com/best-accessible-beauty-products/

Camero, K. (2023, April 20). Rare Beauty’s Products Have Sparked A Critical Conversation About The Beauty Industry’s Lack Of Inclusivity. BuzzFeed News. https://www.buzzfeednews.com/article/katiecamero/rare-beauty-makeup-for-people-with-disabilities

Diverse Perspectives: People with Disabilities Fulfilling Your Business Goals. (n.d.). DOL. Retrieved January 9, 2024, from http://www.dol.gov/agencies/odep/publications/fact-sheets/diverse-perspectives-people-with-disabilities-fulfilling-your-business-goals

Entwistle, V. A., Carter, S. M., Cribb, A., & McCaffery, K. (2010). Supporting Patient Autonomy: The Importance of Clinician-patient Relationships. Journal of General Internal Medicine, 25(7), 741–745. 10.1007/s11606-010-1292-2

Factsheet on Persons with Disabilities | United Nations Enable. (n.d.). Retrieved January 9, 2024, from https://www.un.org/development/desa/disabilities/resources/factsheet-on-persons-with-disabilities.html

Gallagher, G. (2021, March 25). How the Beauty World Can Cater to People with Disabilities. Sunday Edit. https://edit.sundayriley.com/how-the-beauty-world-can-cater-to-people-with-disabilities/

Gregg, N. (2023, February 9). Brands Are Making More Accessible Beauty Products – But Is It Enough? The Zoe Report. https://www.thezoereport.com/beauty/accessibility-in-the-beauty-industry

HOW ACCESSIBLE PACKAGING IMPACT THE EXPERIENCE OF PEOPLE WITH DISABILITIES. (2022, June 2). Blog Indumak - Packers, Balers and Palletizing. https://blog.indumak.com.br/en/how-accessible-packaging-impact-the-experience-of-people-with-disabilities/

Kim, D. (2016). The effects of hand strength on upper extremity function and activities of daily living in stroke patients, with a focus on right hemiplegia. Journal of Physical Therapy Science, 28(9), 2565–2567. 10.1589/jpts.28.2565

Kim, S. (2022, November 16). The beauty industry might actually be catching up on accessibility. Mic. https://www.mic.com/identity/accessible-beauty-products-disability-friendly

Mano, H., Fujiwara, S., & Haga, N. (2018). Adaptive behaviour and motor skills in children with upper limb deficiency. Prosthetics and Orthotics International, 42(2), 236–240. 10.1177/0309364617718411

Nast, C. (2021, June 30). Beauty weak spot: People with disabilities. Vogue Business. https://www.voguebusiness.com/beauty/beauty-fails-people-with-disabilities-loreal-estee-lauder-unilever-wants-to-change-that

Pollock, A., Farmer, S. E., Brady, M. C., Langhorne, P., Mead, G. E., Mehrholz, J., & van Wijck, F. (2014). Interventions for improving upper limb function after stroke. The Cochrane Database of Systematic Reviews, 2014(11), CD010820. 10.1002/14651858.CD010820.pub2

Richards, L., & Pohl, P. (1999). Therapeutic Interventions to Improve Upper Extremity Recovery and Function. Clinics in Geriatric Medicine, 15(4), 819–832. 10.1016/S0749-0690(18)30033-8

Synek, S. S., Lohman, H., & Jewell, V. (2023). Effectiveness of Upper Extremity Orthotic Interventions on Functional Participation for Adults With Stroke: A Systematic Review. The American Journal of Occupational Therapy, 77(Supplement_2), 7711510299p1. 10.5014/ajot.2023.77S2-PO299

Toscano, C. (2023a). Rare Beauty by Selena Gomez is makeup that’s easy to use one-handed— Reviewed. https://reviewed.usatoday.com/accessibility/content/rare-beauty-accessible-packaging

Toscano, C. (2023b). Selena Gomez’s makeup packaging makes it accidentally accessible. Reviewed. https://reviewed.usatoday.com/accessibility/content/rare-beauty-accessible-packaging

WHO Snapshot. (n.d.). Retrieved January 9, 2024, from https://www.who.int/news-room/fact-sheets/detail/disability-and-health

Wu, Y.-L., & Chao, S.-R. (2023). The Effects of a Beauty Program on Self-Perception of Aging and Depression among Community-Dwelling Older Adults in an Agricultural Area in Taiwan. *Healthcare (Basel*, Switzerland), 11(10), 1377. 10.3390/healthcare11101377

Yamamoto, H., Takeda, K., Koyama, S., Morishima, K., Hirakawa, Y., Motoya, I., Sakurai, H., Kanada, Y., Kawamura, N., Kawamura, M., & Tanabe, S. (2022). The relationship between upper limb function and activities of daily living without the effects of lower limb function: A cross-sectional study. British Journal of Occupational Therapy, 85(5), 360–366. 10.1177/03080226211030088

Yoo, C., & Park, J. (2015). Impact of task-oriented training on hand function and activities of daily living after stroke. Journal of Physical Therapy Science, 27(8), 2529–2531. 10.1589/jpts.27.2529

Zhang, L., Adique, A., Sarkar, P., Shenai, V., Sampath, M., Lai, R., Qi, J., Wang, M., & Farage, M. A. (2020). The Impact of Routine Skin Care on the Quality of Life. Cosmetics, 7(3), Article 3. 10.3390/cosmetics7030059

